# Antibody responses to endemic coronaviruses modulate COVID-19 convalescent plasma functionality

**DOI:** 10.1101/2020.12.16.20248294

**Authors:** William Morgenlander, Stephanie Henson, Daniel Monaco, Athena Chen, Kirsten Littlefield, Evan M. Bloch, Eric Fujimura, Ingo Ruczinski, Andrew R. Crowley, Harini Natarajan, Savannah E. Butler, Joshua A. Weiner, Mamie Z. Li, Tania S. Bonny, Sarah E. Benner, Ashwin Balagopal, David Sullivan, Shmuel Shoham, Thomas C. Quinn, Susan Eshleman, Arturo Casadevall, Andrew D. Redd, Oliver Laeyendecker, Margaret E. Ackerman, Andrew Pekosz, Stephen J. Elledge, Matthew Robinson, Aaron A.R. Tobian, H. Benjamin Larman

**Affiliations:** Institute for Cell Engineering, Division of Immunology, Department of Pathology, Johns Hopkins University School of Medicine, Baltimore, MD, USA; Department of Biostatistics, Johns Hopkins University Bloomberg School of Public Health, Baltimore, MD, USA; W. Harry Feinstone Department of Molecular Microbiology and Immunology, Johns Hopkins University Bloomberg School of Public Health, Baltimore, MD, USA; Division of Transfusion Medicine, Department of Pathology, Johns Hopkins University School of Medicine, Baltimore, MD, USA; Division of Genetics, Department of Medicine, Howard Hughes Medical Institute, Brigham and Women’s Hospital, Boston, MA, USA; Department of Genetics, Program in Virology, Harvard University Medical School, Boston, MA, USA; Department of Microbiology and Immunology, Geisel School of Medicine at Dartmouth, Dartmouth College, Hanover, NH, USA; Thayer School of Engineering, Dartmouth College, Hanover, NH, USA; Division of Infectious Diseases, Department of Medicine, Johns Hopkins University School of Medicine, Baltimore, MD, USA; Division of Intramural Research, National Institute of Allergy and Infectious Diseases, National Institutes of Health, Bethesda, MD, USA

## Abstract

COVID-19 convalescent plasma, particularly plasma with high-titer SARS-CoV-2 (CoV2) antibodies, has been successfully used for treatment of COVID-19. The functionality of convalescent plasma varies greatly, but the association of antibody epitope specificities with plasma functionality remains uncharacterized. We assessed antibody functionality and reactivities to peptides across the CoV2 and the four endemic human coronavirus (HCoV) genomes in 126 COVID-19 convalescent plasma donations. We found strong correlation between plasma functionality and polyclonal antibody targeting of CoV2 spike protein peptides. Antibody reactivity to many HCoV spike peptides also displayed strong correlation with plasma functionality, including pan-coronavirus cross-reactive epitopes located in a conserved region of the fusion peptide. After accounting for antibody cross-reactivity, we identified an association between greater alphacoronavirus NL63 antibody responses and development of highly neutralizing antibodies to SARS-CoV-2. We also found that plasma preferentially reactive to the CoV2 receptor binding domain (RBD), versus the betacoronavirus HKU1 RBD, had higher neutralizing titer. Finally, we developed a two-peptide serosignature that identifies plasma donations with high anti-S titer but that suffer from low neutralizing activity. These results suggest that analysis of coronavirus antibody fine specificities may be useful for selecting therapeutic plasma with desired functionalities.

## Introduction

Coronaviruses cause human respiratory diseases that range from asymptomatic to fatal. Endemic human coronaviruses (HCoVs) that cause the common cold include two alphacoronaviruses (229E and NL63) and two betacoronaviruses (OC43 and HKU1). Middle Eastern Respiratory Syndrome (MERS) coronavirus and the Severe Acute Respiratory Syndrome coronavirus (SARS-CoV-1) are betacoronaviruses that cause severe pneumonia. In late 2019, a novel severe pneumonia-causing betacoronavirus, SARS-CoV-2 (CoV2), was described in Wuhan, China. As of November 2020, the COVID-19 pandemic, caused by the spread of SARS-CoV-2, had infected over 50 million individuals and resulted in over one million deaths worldwide. (1)

There is growing evidence for the potential of anti-CoV2 antibodies to treat COVID-19. CoV2 antibodies have been administered in the form of monoclonal antibodies directed at specific CoV2 epitopes and hyperimmune globulin or convalescent plasma obtained from individuals who recovered from COVID-19. (2-5) Convalescent plasma has received Emergency Use Authorization from the United States Food and Drug Administration for treatment of COVID-19; preliminary data support the efficacy of convalescent plasma, specifically for donations that contain high titers of CoV2 antibodies. (5)

The antibody response to CoV2 is highly variable in terms of titer, (6, 7) avidity, (8) antigenic preference, (9, 10) kinetics of induction, (7) isotype usage, (11) and functionally protective capacity. (11) Differential pre-existing immune responses to endemic HCoVs may contribute to the large variation in CoV2 antibody response. Recent studies of pre-pandemic plasma identified a low prevalence of pre-existing reactivity against the S2 subunit of the CoV2 S. (11) S2 contains structures that are critical for virus entry into cells, such as the fusion peptide, which is conserved across coronaviruses; sequence conservation in this region may explain the presence of these CoV2-reactive antibodies prior to the COVID-19 pandemic. (10) Boosting of pre-existing HCoV antibodies in response to infection with CoV2 may occur in the absence of antibody functionality, a phenomenon referred to as “original antigenic sin.” (12) Alternatively, HCoV antibody responses may prove beneficial during CoV2 infection, as HCoV neutralization activity has been correlated with decreased disease severity, (13) and anti-CoV2 activity of preexisting HCoV antibodies has been suggested. (14) There remains an important gap in understanding the relationships between cross-reactive HCoV antibodies, convalescent plasma CoV2 antibody binding specificities, and the functional activities of COVID-19 convalescent plasma.

In this study, we used systems serology and massively multiplexed epitope profiling to characterize the functionality and fine specificities of coronavirus antibodies in a cohort of convalescent COVID-19 plasma donors (Fig 1a). We correlated dominant CoV2 and HCoV peptide reactivities with viral neutralization, antibody dependent cellular phagocytosis (ADCP), antibody dependent cellular cytotoxicity (ADCC), and antibody dependent complement deposition (ADCD). We developed an algorithm to deconvolute cross-reactivity among homologous peptides, which helped explain how disparate HCoV antibody responses may modulate functional characteristics of CoV2 convalescent plasma.

**Figure 1.**
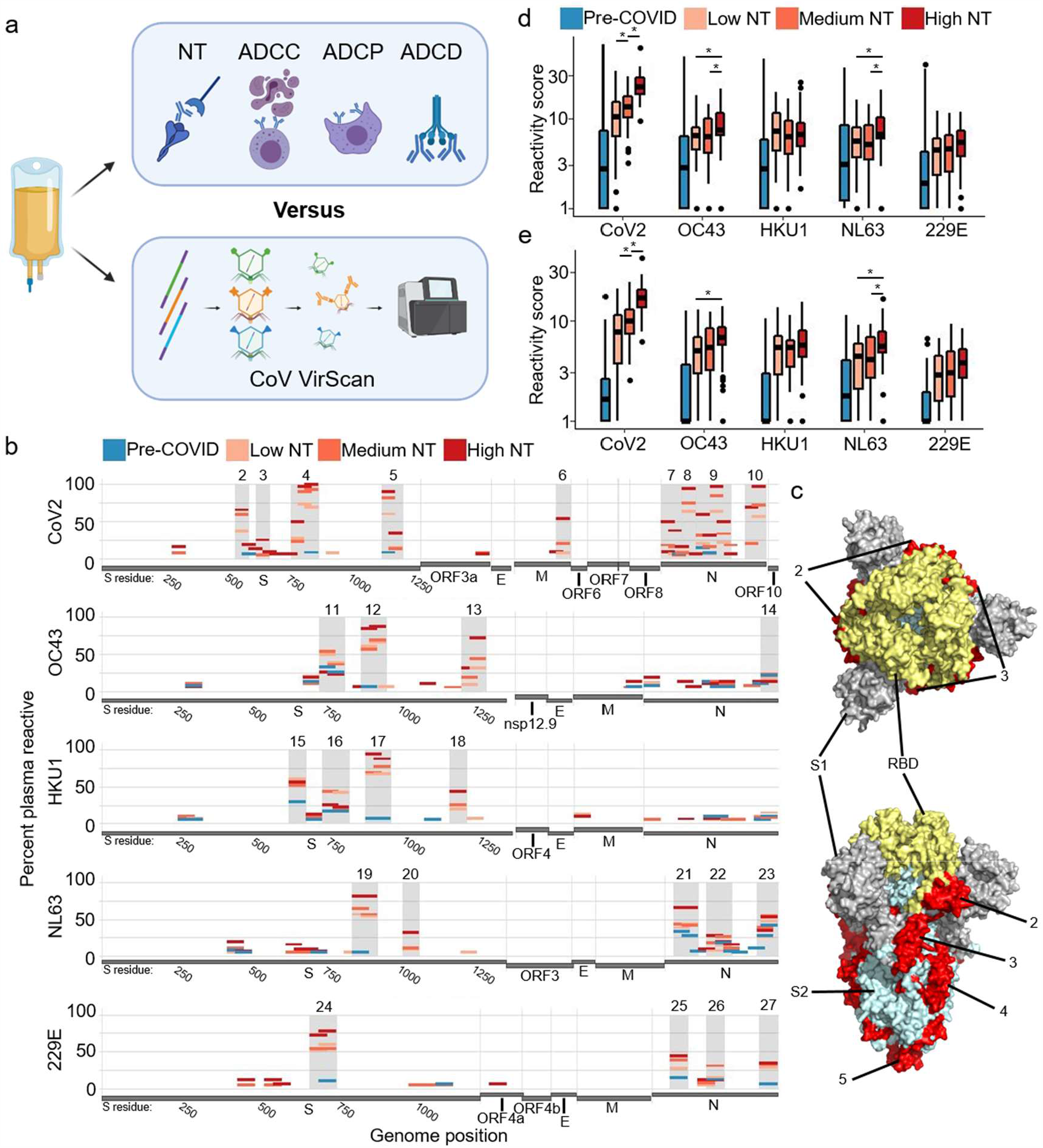
Correlating coronavirus peptide reactivity and neutralizing titer of COVID-19 convalescent plasma. **a**. 126 COVID-19 convalescent plasma donations underwent functional analysis and antibody profiling via VirScan with a comprehensive coronavirus peptide library. Functionalities included neutralizing titer (NT), antibody dependent cellular cytotoxicity (ADCC), antibody dependent cellular phagocytosis (ADCP), and antibody dependent complement deposition (ADCD). Plasma from 87 pre-pandemic controls were additionally analyzed via VirScan. **b**. Convalescent plasma was divided into groups based on NT AUC (Low NT: <40 – n=55, Medium NT: 40 to 160 – n=39, and High NT: ≥160 – n=32). The percentages of samples in each group with reactivity to a particular peptide were plotted according to the peptide’s position along each viral genome. Regions with most consistent reactivity were shaded and were located in S and N. Amino acid residue number is included for S. Full genome plots are shown in Figure S1. **c**. Immunodominant regions of CoV2 S are mapped onto the CoV2 S structure. (31) Trimeric spike is colored by S1 subunit and S2 subunit with the entire receptor binding domain (RBD) indicated. **d**. Aggregate virus score was calculated as the sum of all log-transformed fold changes of peptides designed for a given virus. Bars with an asterisk indicate convalescent plasma groups that had significantly different scores (two sided Wilcox test, p<0.05) **e**. Aggregate virus scores from peptides defining dominant regions.

## Results

### Systems serology and VirScan analysis of convalescent plasma donations

Convalescent plasma for this study were provided by 126 donors for therapeutic use 13-67 days after PCR-confirmed CoV2 infection. (6, 8) As part of a therapeutic plasma treatment trial, CoV2 neutralizing titer (NT) was measured for each sample and reported as an area under the curve (NT AUC). We divided convalescent plasma donations into groups according to their neutralizing activity: High NT (NT AUC ≥160, n=32), Medium NT (NT AUC ≥40 and <160, n=39), and Low NT (NT AUC <40, n=55). (6) We also measured ADCP, ADCC, and ADCD for each plasma donation. (11) We recently used a massively multiplexed antibody profiling system (VirScan) to analyze peptide epitopes across over 230 COVID-19 and 190 Pre-COVID samples. (10) This library contains 3,466 peptides, 56 amino acids in length, that tile across the CoV2 and all four endemic HCoV genomes with 28 amino acid overlaps; the library also covers SARS-CoV-1, MERS, and three related bat CoVs: BatCoV-Rp3, BatCoV-HKU3 and BatCoV-279. (10) At several genomic locations, single amino acid variants of CoV2 are also represented in the library. We used this coronavirus VirScan library to identify the epitopes targeted in each of the 126 plasma donations. For comparison, we also performed coronavirus VirScan on 87 Pre COVID-19 (Pre-COVID) pandemic plasma samples. (15)

In all plasma samples, we measured antibody binding to each 56 amino acid peptide in the library (Fig 1b, Fig S1). Using a threshold of 20% reactivity in any sample group, we identified 27 regions of frequent (dominant) reactivity that largely corresponded to regions we had previously established. (10) Among the samples that harbored dominant reactivities, the magnitude of reactivity to specific peptides was not associated with NT AUC, with one exception: reactivity to an immunodominant region that overlaps the CoV2 S RBD (residues 533-588) was greater in High NT plasma (Fig S2, Wilcox test: vs moderate p=0.014, vs poor p<0.001). At the peptide level, it therefore appears that the response clonality, defined by the number of reactive epitopes, rather than the magnitude of reactivity to individual peptides, distinguishes Low NT from High NT plasma.

Table 1 provides the genomic location and reactivity profiles for each immunodominant peptide (amino acid sequences in Table S1). Peptides with identical start and stop locations represent genomic variants. Three regions of CoV2 S, one region of the CoV2 membrane protein (M), and four regions of the CoV2 nucleocapsid protein (N) were recognized by 50-100% of High NT plasma. Antibodies in Medium NT and Low NT plasma recognized many of the same regions, but at a lower frequency. Exceptions to this pattern were that Low NT plasma displayed the most frequent reactivity to peptides around the S1/S2 cleavage site (CS) from the betacoronaviruses HKU1 and OC43. Many HCoV dominant regions showed increasing frequency of reactivity with increasing NT AUC, including the highly conserved S fusion peptide (FP), which is required for cell entry of coronaviruses. We previously identified the CoV2 FP as one of the most reactive public CoV2 epitopes; the corresponding peptide from HCoV viruses show high conservation and boosting in CoV2 patients. (10) Peptide reactivities associated with NT AUC frequently correlated with other plasma functionalities (Table 1); this was not surprising, since these functionalities were all independently correlated with NT AUC. (11) Interestingly, the peptide reactivity that had the strongest correlation with ADCC derives from an intra-virion portion of CoV2 M (residues 169-224) which is a known T cell epitope. (16) For structural context, we mapped the dominant CoV2 S regions onto the mature folded S protein structure (Fig 1c). (17)

**Table 1.** Immunodominant coronavirus peptides and their functional correlates. 52 peptides defining 27 immunodominant regions were identified. The percentage of samples with a specific reactivity is shown (red shading). Associations with plasma functionality were defined by dichotomizing all plasma as positive or negative for a particular reactivity, followed by a two sided Wilcox test. The negative log transformed p values are shown (green shading). Adjusted significance cutoff was 0.0255 or, negative log transformed, 1.59 (Benjamini-Hochberg).

|  | Virus | Segment | Domain<br>(Spike Only) | Start | End | Percent Samples Enriched |  |  |  | Association with Phenotype (-log(p)) |  |  |  |
| --- | --- | --- | --- | --- | --- | --- | --- | --- | --- | --- | --- | --- | --- |
|  |  |  |  |  |  | Pre-COVID<br>n=87 | Low NT<br>n=55 | Medium<br>NT<br>n=39 | High NT<br>n=32 | NT AUC | ADCP | ADCC | ADCD |
| 1 | SARS-CoV-2 | ORF1 |  | 281 | 336 | 19.5 | 23.6 | 23.1 | 21.9 | 0.1 | 0.1 | 0.3 | 0 |
| 2 | SARS-CoV-2 | S | RBD | 533 | 588 | 2.3 | 36.4 | 59 | 65.6 | 2.8 | 5 | 3.6 | 1.1 |
| 3 | SARS-CoV-2 | S | CS | 617 | 672 | 1.1 | 0 | 5.1 | 25 | 3.5 | 4.5 | 3.6 | 1 |
| 4 | SARS-CoV-2 | S | FP | 757 | 812 | 0 | 27.3 | 23.1 | 50 | 1.8 | 2.8 | 1.9 | 3.1 |
|  |  |  |  | 757 | 812 | 0 | 20 | 17.9 | 34.4 | 1 | 2.4 | 2.3 | 3.1 |
|  |  |  |  | 785 | 840 | 4.6 | 67.3 | 79.5 | 96.9 | 3.1 | 3.6 | 3.8 | 3.3 |
|  |  |  |  | 785 | 840 | 3.4 | 72.7 | 89.7 | 96.9 | 3.5 | 3.6 | 4 | 3 |
|  |  |  |  | 813 | 868 | 8 | 69.1 | 92.3 | 100 | 4.5 | 3.9 | 3.2 | 2.2 |
| 5 | SARS-CoV-2 | S | HR2 | 1121 | 1176 | 8 | 60 | 82.1 | 90.6 | 3 | 3 | 1.3 | 1.4 |
|  |  |  |  | 1149 | 1204 | 3.4 | 14.5 | 12.8 | 34.4 | 1.3 | 1.6 | 1.8 | 1.5 |
| 6 | SARS-CoV-2 | M |  | 169 | 224 | 0 | 7.3 | 20.5 | 53.1 | 5.3 | 6.4 | 7.4 | 1.8 |
| 7 | SARS-CoV-2 | N |  | 1 | 56 | 1.1 | 9.1 | 17.9 | 50 | 4.8 | 4.7 | 6.4 | 3.5 |
|  |  |  |  | 29 | 84 | 0 | 5.5 | 15.4 | 37.5 | 4.4 | 3.5 | 4.4 | 2.1 |
| 8 | SARS-CoV-2 | N |  | 85 | 140 | 1.1 | 63.6 | 74.4 | 93.8 | 3.7 | 5.3 | 2.8 | 1.7 |
| 9 | SARS-CoV-2 | N |  | 141 | 196 | 13.8 | 18.2 | 25.6 | 46.9 | 1.8 | 1.3 | 3.6 | 3.2 |
|  |  |  |  | 141 | 196 | 14.9 | 20 | 30.8 | 59.4 | 2.5 | 1.9 | 3.2 | 3.3 |
|  |  |  |  | 169 | 224 | 2.3 | 5.5 | 15.4 | 31.3 | 2.9 | 0.9 | 3 | 3.4 |
|  |  |  |  | 169 | 224 | 6.9 | 3.6 | 10.3 | 28.1 | 2.8 | 1.6 | 2.4 | 2.9 |
|  |  |  |  | 197 | 252 | 2.3 | 49.1 | 76.9 | 96.9 | 5 | 4.9 | 2.5 | 2.2 |
|  |  |  |  | 197 | 252 | 3.4 | 63.6 | 84.6 | 96.9 | 7.1 | 6.8 | 3.7 | 2.4 |
|  |  |  |  | 225 | 280 | 1.1 | 21.8 | 33.3 | 50 | 2.1 | 3.5 | 4.4 | 2.3 |
| 10 | SARS-CoV-2 | N |  | 337 | 392 | 2.3 | 20 | 20.5 | 68.8 | 4.5 | 4.2 | 4.3 | 2.7 |
|  |  |  |  | 337 | 392 | 2.3 | 18.2 | 20.5 | 59.4 | 5.2 | 5.2 | 5.5 | 2.9 |
|  |  |  |  | 365 | 420 | 3.4 | 56.4 | 71.8 | 96.9 | 4.3 | 4.3 | 4.1 | 0.7 |
| 11 | OC43 | S | CS | 729 | 784 | 32.2 | 49.1 | 53.8 | 25 | 1.3 | 1.2 | 1.1 | 0.2 |
|  |  |  |  | 757 | 812 | 25.3 | 38.2 | 35.9 | 21.9 | 0.8 | 0.7 | 0.3 | 0.2 |
| 12 | OC43 | S | FP | 869 | 924 | 5.7 | 60 | 56.4 | 84.4 | 1.2 | 1.6 | 2.8 | 0.5 |
|  |  |  |  | 897 | 952 | 4.6 | 65.5 | 69.2 | 87.5 | 1.7 | 1.7 | 2.5 | 0.9 |
| 13 | OC43 | S | HR2 | 1205 | 1260 | 2.3 | 9.1 | 17.9 | 31.3 | 1.7 | 1.8 | 0.9 | 1.1 |
|  |  |  |  | 1233 | 1288 | 1.1 | 30.9 | 43.6 | 71.9 | 2.8 | 2 | 1.9 | 1.9 |
| 14 | OC43 | N |  | 393 | 448 | 20.7 | 16.4 | 20.5 | 12.5 | 0.4 | 0.3 | 0.2 | 0.8 |
|  |  |  |  | 394 | 449 | 20.7 | 18.2 | 23.1 | 12.5 | 0.5 | 0.4 | 0.1 | 0.1 |
| 15 | HKU1 | S | RBD | 617 | 672 | 28.7 | 60 | 51.3 | 56.3 | 0.3 | 0.3 | 0.3 | 0.5 |
| 16 | HKU1 | S | CS | 729 | 784 | 17.2 | 43.6 | 43.6 | 25 | 0.7 | 0.2 | 0.3 | 0.2 |
|  |  |  |  | 757 | 812 | 17.2 | 41.8 | 20.5 | 21.9 | 1.3 | 0.4 | 0.1 | 0.3 |
| 17 | HKU1 | S | FP | 869 | 924 | 6.9 | 67.3 | 69.2 | 93.8 | 1.9 | 2.4 | 2.5 | 0.9 |
|  |  |  |  | 897 | 952 | 6.9 | 67.3 | 76.9 | 87.5 | 1.2 | 1.5 | 1.8 | 0.9 |
| 18 | HKU1 | S | HR2 | 1149 | 1204 | 0 | 20 | 25.6 | 43.8 | 1.8 | 2.8 | 3.6 | 4.7 |
| 19 | NL63 | S | FP | 841 | 896 | 5.7 | 56.4 | 64.1 | 81.3 | 1.9 | 1.6 | 1.4 | 0.8 |
|  |  |  |  | 869 | 924 | 3.4 | 54.5 | 56.4 | 81.3 | 1.9 | 1.5 | 1.7 | 1.1 |
| 20 | NL63 | S | HR1 | 1009 | 1064 | 1.1 | 9.1 | 10.3 | 31.3 | 2.1 | 1.1 | 0.9 | 0.8 |
| 21 | NL63 | N |  | 29 | 84 | 33.3 | 38.2 | 43.6 | 65.6 | 0.8 | 0.1 | 0.2 | 0.3 |
|  |  |  |  | 57 | 112 | 27.6 | 43.6 | 41 | 65.6 | 0.7 | 0 | 0.2 | 0.2 |
| 22 | NL63 | N |  | 141 | 196 | 3.4 | 7.3 | 17.9 | 28.1 | 2.2 | 1.5 | 1.6 | 1 |
|  |  |  |  | 169 | 224 | 18.4 | 14.5 | 25.6 | 18.8 | 0.8 | 0.5 | 0.1 | 0.1 |
| 23 | NL63 | N |  | 309 | 364 | 27.6 | 25.5 | 38.5 | 34.4 | 0.3 | 0.3 | 0.2 | 0.1 |
|  |  |  |  | 323 | 378 | 41.4 | 47.3 | 53.8 | 53.1 | 0.1 | 0.4 | 0.3 | 0.3 |
| 24 | 229E | S | FP | 645 | 700 | 2.3 | 52.7 | 53.8 | 71.9 | 1.4 | 0.9 | 1.4 | 0.2 |
|  |  |  |  | 673 | 728 | 10.3 | 60 | 53.8 | 78.1 | 1 | 0.8 | 1.4 | 0 |
| 25 | 229E | N |  | 57 | 112 | 14.9 | 27.3 | 38.5 | 43.8 | 0.5 | 0 | 0.2 | 0.7 |
| 26 | 229E | N |  | 169 | 224 | 12.6 | 14.5 | 30.8 | 12.5 | 0.3 | 0.9 | 0.3 | 0.1 |
| 27 | 229E | N |  | 337 | 392 | 6.9 | 29.1 | 33.3 | 34.4 | 0.4 | 0.1 | 0.5 | 0.5 |

To correlate putative pre-existing HCoV-specific immune responses with CoV2-specific responses, we developed two aggregate, virus-level reactivity scores: (i) aggregate reactivity across all peptides from each virus (Fig 1d), and (ii) aggregate reactivity of the dominant peptides from each virus (Fig 1e). The aggregate CoV2 scores were highest in High NT plasma, as expected. High NT plasma also exhibited significantly greater aggregate NL63 and OC43 reactivity compared to Low NT plasma, potentially reflecting some degree of cross-reactivity with CoV2. In addition, a higher number of total reactivities (polyclonality) in the dominant regions of CoV2 and NL63 correlated with higher NT AUC (Fig S3, Pearson’s correlations: CoV2 p<10^−8^, NL63 p=0.02).

### SARS-CoV-2 and endemic coronavirus cross-reactivity

Many immunodominant CoV2 peptides are homologous to HCoV peptides, and HCoV peptides were more frequently recognized by the convalescent plasma compared to Pre-COVID plasma. We therefore evaluated potential cross-reactivity among dominant epitopes. We identified two independent clusters of peptides from CoV2 and the HCoVs that exhibited significantly correlated reactivities (Fig 2a). The largest cluster of highly correlated peptide reactivities was comprised of the fusion peptide (FP) from the S2 subunit of each HCoV S; all of these peptides showed sequence homology to the immunodominant CoV2 FP peptides (Fig 2b). The immunodominant FP peptides were the most frequently recognized peptides from each coronavirus among all plasma NT groups (Table 1). Plasma with antibodies to CoV2 FP most often also reacted with all other CoV FPs, including the zoonotic bat CoVs represented in our library (Fig S4). The smaller cluster of correlated reactivity in Fig2a contained a CoV2 S HR2 peptide and the homologous OC43 S HR2 peptides. Of note, CoV2 FP and HR2 peptides were targeted by antibodies in up to 8% of Pre-COVID plasma. This suggests that the immunodominant CoV2 FP and HR2 regions may be key epitopes of pre-existing HCoV antibodies that may provide heterologous protection from CoV2 infection and/or are boosted during CoV2 antibody responses.

**Figure 2.**
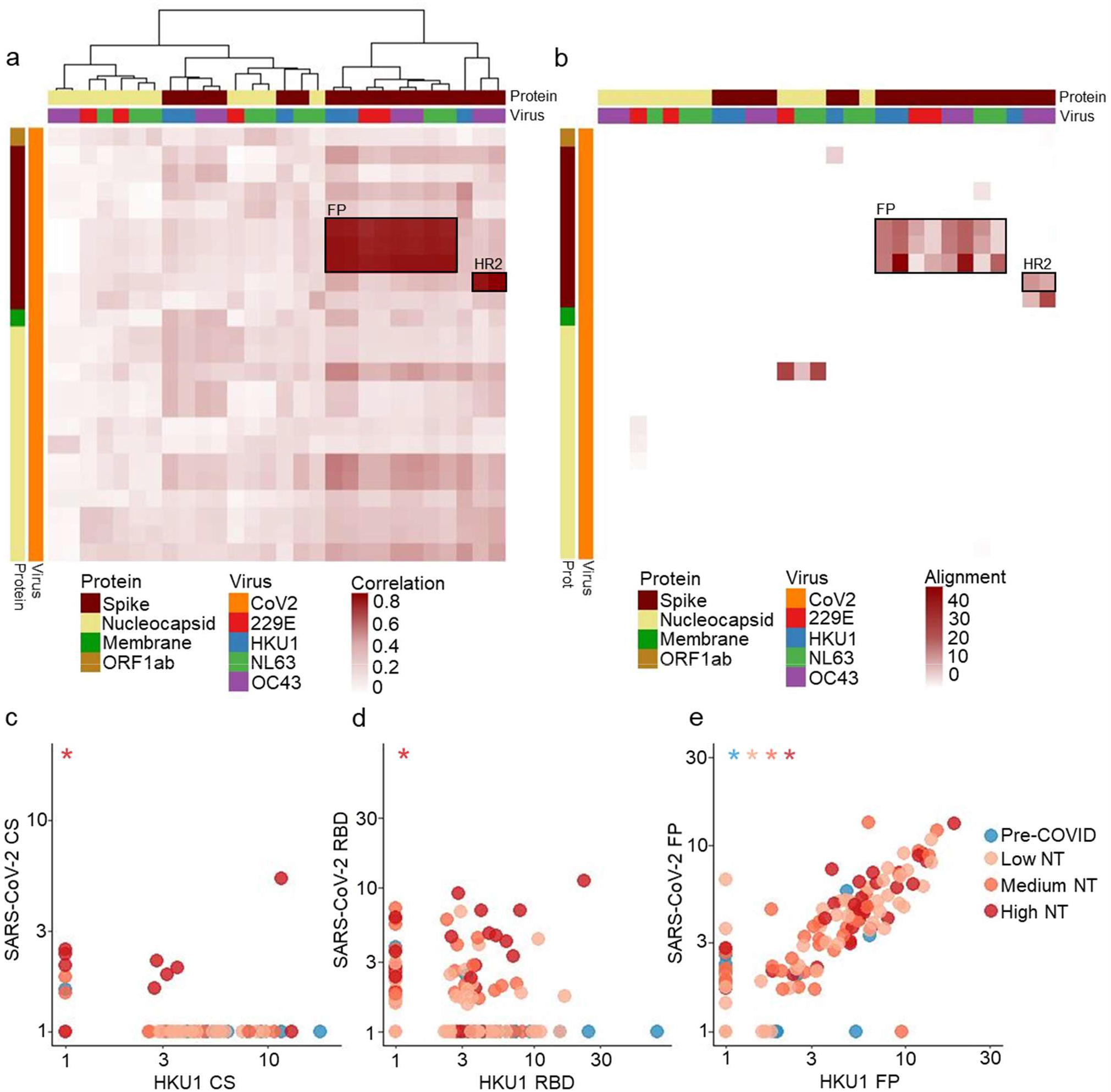
Reactivities against some CoV2 peptides are highly correlated with reactivities against homologous HCoV peptides. **a**. Spearman correlation coefficient matrix between dominant CoV2 peptides and dominant HCoV peptides is shown in the form of a clustered heatmap. CoV2 peptides (y-axis) are ordered by genomic location from top to bottom, while HCoV peptides (x-axis) were clustered according to their correlations. The heatmap annotations depict peptides’ overall frequency of antibody reactivity, protein of origin, and virus of origin. Highly correlated peptides map to Fusion Peptide (FP) or Heptad Repeat 2 (HR2) **b**. Sequence similarity (as defined by the negative log of the blastp evalue) between dominant CoV2 peptides in CoV2 and dominant HCoV peptides is shown. The rows and columns of the heatmap match those of the correlation heatmap to facilitate comparison. The regions of highest correlation (boxes as in **a**) show strongest alignment. **c-e**. Antibody reactivity (measured as fold changes) to three HKU1 S peptides are plotted against reactivity to homologous peptides of CoV2 S. The two CS peptides have no sequence homology and their reactivity is not correlated. The two RBD peptides have moderate homology and show frequent co-reactivity but no strong correlation. The two FP peptides have high sequence homology and strong correlation among all sample groups. Asterisks indicate Pearson correlation with nonzero coefficient for a given plasma group (p<0.05).

We next evaluated the relationships between CoV2 peptides and homologous HCoV peptides by comparing the magnitudes of the reactivities towards the dominant CS, RBD, and FP peptides from CoV2 and HKU1 (Fig 2c-e). The two FP peptides (CoV2 residues 813-868, HKU1 residues 869-924) had significant amino acid level homology and a strong positive correlation in antibody reactivity regardless of sample group (including Pre-COVID). Meanwhile, reactivity to the two CS peptides (CoV2 residues 617-672, HKU1 residues 757-812) that did not share amino acid homology was uncorrelated, except for in highly neutralizing plasma that often reacted to both. The two RBD peptides (CoV2 residues 533-588, HKU1 residues 617-672) had a moderate level of amino acid homology and were frequently co-reactive, but only the High NT plasma had a nonzero correlation between fold changes for CoV2 RBD and HKU1 RBD (Pearson’s correlation, p<0.001).

The CoV2 and HKU1 RBD peptide reactivities were sometimes independently reactive; recognition of RBD peptides from both viruses may reflect cross-reactivity and/or independent, non-cross-reactive (yet correlated) reactivities. These examples highlight the complex interplay between pre-existing, boosted, and de-novo cross-reactive antibody responses to coronaviruses.

### Deconvoluting antibody specificity from VirScan data

We hypothesized that an individual’s HCoV immune response history might impact the spectrum of epitopes targeted during CoV2 infection and the resultant plasma functionality. Clearly, however, many HCoV reactivities observed in the CoV2 convalescent plasma are likely due to cross-reactive CoV2 antibodies as we described previously. (10) We therefore sought to “deconvolute” the VirScan data, assuming that antibodies would exhibit preference for their intended “on-target” peptides compared with cross-reactive peptides (see Methods, Deconvolution, schematic in Fig S5). Deconvoluted profiles retain only definitive target-preferred reactivities.

The deconvoluted VirScan prevalence data is plotted on the coronavirus genomes in Fig 3a. As expected, Pre-COVID CoV2 reactivity largely disappeared, since these samples can only harbor cross-reactive antibody binding to CoV2. Additionally, many CoV2 S reactivities highly correlated with homologous HCoV peptide reactivities, such as the FP reactivities, were reduced due to insufficient support for preference of a specific coronavirus. Some reactivities against the CoV2 S RBD and HR2 were also removed, whereas reactivities around the CoV2 S CS region were retained (Fig 3b, dominant region 3). Nucleocapsid (N) peptides, meanwhile, tended to be more readily distinguishable as non-cross-reactive. Most reactivities against OC43 (the HCoV most closely related to CoV2)(11) were either unattributed or attributed to antibodies targeting CoV2, and were thus removed by deconvolution. Aggregated deconvoluted CoV2 and HCoV reactivities were then assessed for correlation with plasma functionality (Fig 3c-d). As expected, NT AUC remained highly correlated with aggregated deconvoluted CoV2 reactivity, while the OC43 association with NT AUC was no longer supported by deconvoluted data. After deconvolution, the increased NL63 score remained associated with higher NT AUC (Wilcox test, p=0.005), further supporting a role for prior NL63 infection in increased CoV2 NT AUC.

**Figure 3.**
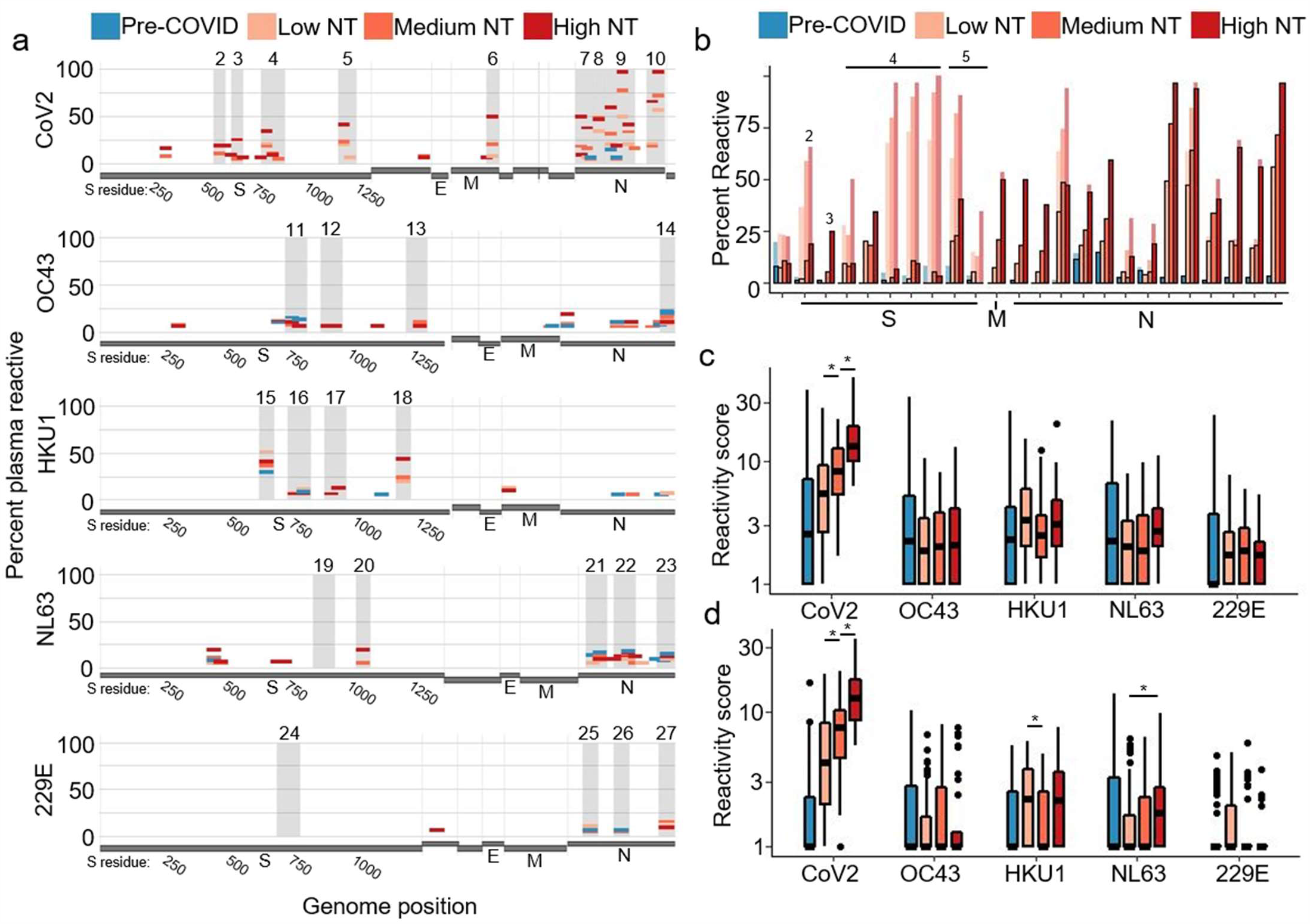
Deconvolution of convalescent plasma reactivities. **a**. Following deconvolution, the percentage of samples in each sample group with target-preferred peptide reactivities were plotted along the viral genomes. Amino acid residue number is included for S. **b**. The percentage of plasma in each sample group that had reactivity to dominant CoV2 peptides is shown before (light bars) and after (dark bars with outline) deconvolution. **c-d**. Aggregate virus scores were calculated following deconvolution, using all peptides (**c**) or using only peptides from immunodominant regions (**d**). Bars with an asterisk indicate convalescent plasma groups that show significantly different scores (two sided wilcox test, p<0.05)

We next evaluated associations between individual deconvoluted peptide level reactivities with plasma functionality (Table S2). The most immunodominant CoV2 S RBD peptide (residues 533-588) was preferentially targeted in 18.8% of High NT plasma compared to 10.3% of Medium NT plasma, and only 1.8% of Low NT plasma (Fisher’s, High NT vs Low NT p=0.009). In contrast, HKU1-preferred RBD (residues 617-672) antibodies were identified in 40.6% of High NT plasma and 50.9% of Low NT plasma (difference not significant). These results indicate that CoV2-preferred antibody reactivity to RBD, versus HKU1-preferred antibody reactivity to RBD, is associated with higher NT AUC.

### Discordance of CoV2 spike antibody titer versus NT AUC

The suitability of plasma donations is typically evaluated by measuring total S antibody titer using an enzyme linked immunosorbent assay (ELISA). Total S antibodies correlate with neutralization in our cohort and others. (6, 7, 18) However, S antibodies tend to be more sensitive than specific for neutralization. Using VirScan, we investigated whether reactivities to specific peptides were associated with discordance between functionality and total S reactivity.

For this analysis, we performed a simple linear regression to predict NT AUC using total S titer determined by a multiplex microsphere assay. (11) Convalescent plasma samples were then categorized into three groups: High NT/Low S, Concordant NT/S, and Low NT/High S (Fig 4a, Methods). To evaluate whether boosting of non-neutralizing HCoV antibodies contributed to lower NT/S ratios, we first tested for associations between aggregate coronavirus scores and NT/S discordance (Fig 4b-c). We found a correlation between aggregate HKU1 score and Low NT/High S (pre-deconvolution Wilcox, p=0.019; post-deconvolution Wilcox, p=0.011). We then explored whether reactivity to specific HKU1 peptides could account for this association. A single HKU1 peptide, the S1/S2 cleavage site peptide (CS, residues 757-812, dominant region 16 in Table 1), was significantly associated with Low NT/High S (Wilcox, p=0.023). The association between HKU1 S CS reactivity and over-prediction of NT AUC depended on the absence of CoV2 CS (residues 617-672) reactivity (Fig 4d). When reactivities to HKU1 CS and CoV2 CS peptides were taken into account, roughly half of the samples with low NT/S were identified as less suitable for transfusion (Fig 4e, Methods). These results establish that the fine specificities of CoV antibodies, including HCoV reactivities, may prove helpful in improving the assessment of plasma functionality for clinical use.

**Figure 4.**
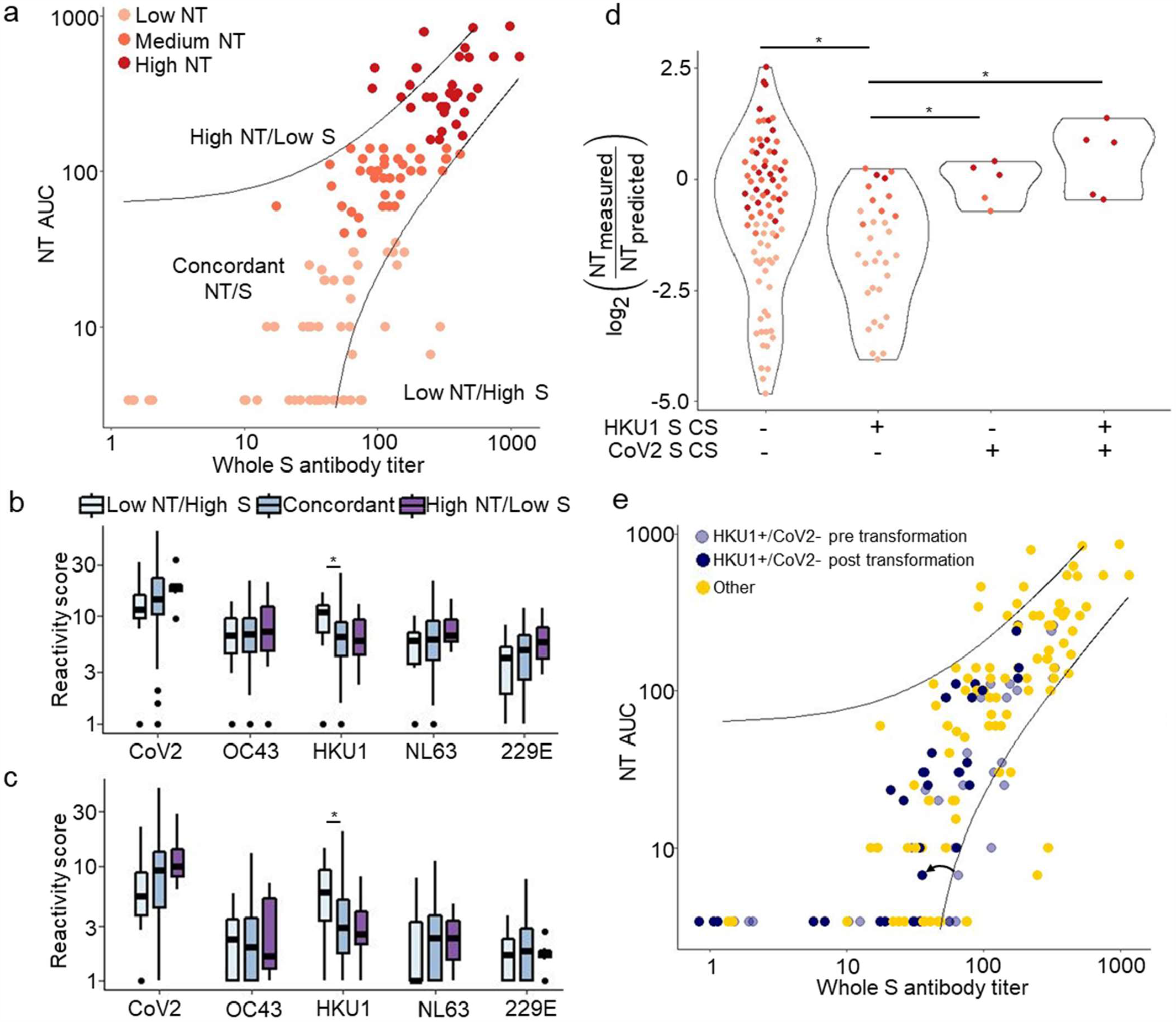
VirScan identifies features associated with discordance between whole spike titer and NT AUC. **a**. NT AUC was plotted against whole S antibody titer. A linear regression was performed between whole S and NT AUC to establish a predicted NT AUC; guidelines were plotted to indicate samples displaying large discordance between NT AUC and whole S titer (Methods). **b-c**. Aggregate virus scores were calculated for plasma donations with concordant NT/S, discordantly Low NT/S, and discordantly High NT/S using pre (**b**) and post (**c**) deconvolution peptide reactivity. Bars indicate differences in aggregate scores between groups (Wilcox Rank Sum, p<0.05) **d**. Ratio of measured NT AUC to predicted NT AUC, versus CoV2 CS and HKU1 CS reactivities. Bars with an asterisk indicate plasma groups that show significantly different scores (Two sided wilcox test, p<0.05) **e**. Plasma defined by the HKU1 CS+/CoV2 CS-reactivity pattern are shown on the scatter plot in **a**. A correction factor (example indicated by arrow) was applied to these plasma to account for the association with NT/S discordance.

## Discussion

In this study, we used VirScan to quantify the relationship between the fine specificities of anti-CoV antibodies and the functionality of COVID-19 convalescent plasma. These analyses allowed us to define peptide epitopes targeted by pre-existing antibodies that cross-react with CoV2. Many antibody reactivities to HCoV peptides observed in the convalescent phase of CoV2 infection likely reflect an anamnestic response (boosting of pre-existing antibody reactivities by CoV2) or a feature of the cross-reactive CoV2 antibody response as previously described. (10) In either case, HCoV reactivities represent potential biomarkers for identifying plasma with differing therapeutic potency. In particular, polyclonal responses to CoV2 and NL63, as well as a cross-reactive response to the conserved fusion peptide (FP) of CoV2 S, were all associated with increased plasma functionality. In contrast, plasma with antibodies that recognize HKU1 CS peptides but not CoV2 CS peptides suffered from relatively low neutralizing activity.

The defining feature of highly neutralizing plasma was high polyclonality of CoV2 antibodies. High NT plasma reacted with most of the same immunodominant epitopes as the low NT plasma, and at relatively similar levels of reactivity, but at much higher frequency. NT, ADCC, ADCP and ADCD have been found to correlate with one another, (11) but individual peptide epitopes displayed variable associations with different, sometimes unexpected, functionalities. It is important to recognize that antibody reactivity to individual peptides may track with increased plasma functionality for two opposing reasons. First, they may be positively correlated with disease severity (since disease severity is positively correlated with convalescent plasma functionality). Alternatively, reactivities may directly contribute to a more effective humoral response (thus tending to negatively correlate with disease severity). Antibodies that react to the immunodominant RBD peptide, FP peptides, and the CS peptide are the most likely antibodies described in this study to be neutralizing. Support for the functional relevance of CS peptide-specific antibodies derives from escape mutations in this region that have arisen in response to therapeutic antibodies. (19) However, the relationship between reactivity to the CS region and disease severity is not known. Beyond CoV2 antibody specificities, aggregation of post-deconvolution NL63 peptide reactivities suggested that a stronger immune response to NL63 is associated with generating highly neutralizing plasma. This result is consistent with the previously reported correlation between NL63 neutralization and milder disease severity, (13) potentially attributable to this virus’s usage of the ACE2 receptor, as is the case for CoV2.

Pre-existing antibodies that react with CoV2 S2 have been described; however, data on the activity of these antibodies is conflicting. (14, 20) We had previously shown that reactivity to the highly conserved FP region might explain pre-existing antibody reactivity to CoV2 S2 and that FP represents a pan-CoV conserved antibody epitope. (10) In this study, a small minority of Pre-COVID plasma were found to contain antibodies reactive to CoV2 FP peptides (≤10%, similar to CoV2 HR2 reactivity). Deconvolution attributed almost all of the observed Pre-COVID reactivity against CoV2 FP and HR2 to cross-reactivity from antibodies that recognize HCoV peptides. The cross-reactive response to HCoV FP and CoV2 FP appears to be boosted by CoV2 infection, and while support for or against the functionality of these antibodies is currently lacking, their correlation with increased serum functionality is consistent with a protective role. If anti-FP antibodies directly confer protection, banked plasma containing anti-FP antibodies may prove useful in future CoV outbreaks (including zoonotic CoVs related to the bat CoVs in our library) and FP would be a candidate antigen for universal CoV vaccination.

Therapeutic use of convalescent plasma with a higher titer of anti-CoV2 antibodies is more likely to be efficacious. (21) As trials of convalescent plasma for prophylactic and/or early therapeutic use show efficacy, demand for convalescent plasma is significantly increasing. Although the FDA initially recommended measuring neutralizing titers for convalescent plasma, the low capacity of BSL-3 laboratories nationwide limits the ability to quantify viral neutralization at the scale required for widespread use. Surrogate assays that measure neutralizing antibodies without requiring a BSL-3 lab are in development, (22) but ELISA-based measurements of antibodies directed against S, S1 or RBD are currently used. We evaluated the specific peptide reactivities associated with the greatest relative difference between whole S antibody titer and neutralizing titer. HKU1 reactivity, specifically directed against the CS peptide, was associated with relatively low functionality. Evaluating convalescent plasma for reactivity to HKU1 S CS and CoV2 S CS may therefore be used to identify plasma most likely to have clinical benefit.

We identified specific HCoV antibody responses that are likely to impact the potency of COVID-19 convalescent plasma. We additionally developed a method using VirScan data to distinguish preferential antibody recognition from ambiguous cross-reactivity, which may be used more broadly for other viruses to better understand original antigenic sin versus heterologous protection. This approach may also be used to augment current VirScan analysis strategies that seek to differentiate antibody responses among related viruses. (23) An important limitation of VirScan is that the phage do not display highly conformational, discontinuous, or glycosylated epitopes. Some, if not most, of the antibody reactivities identified in this study do not confer functional activity to the plasma. However, these reactivities may track closely with functional antibody reactivities that are not detectable by VirScan. Regardless of whether HCoV peptide reactivities are pre-existing or arise in the context of CoV2 infection, we found clear differences in HCoV reactivity between poorly and highly functional plasma. For example, highly functional plasma contained a stronger polyclonal antibody response to NL63. Additionally, HKU1 S CS reactivity confounds prediction of NT AUC, while antibody preference for CoV2 RBD over HKU1 RBD identifies highly active plasma. An understanding of the fine specificities of anti-coronavirus antibody repertoires may be applied to therapeutic plasma prioritization and may serve to generate novel hypotheses regarding the molecular mechanisms underlying the complex immune responses elicited by COVID-19 infection.

## Methods

### Study participants

Candidate convalescent plasma donors were contacted by study personnel, as previously described. (6) All donors were at least 18 years old and had a confirmed diagnosis of SARS-CoV-2 by detection of RNA in a nasopharyngeal swab sample. In addition, donors were informed that they would need to satisfy standard eligibility criteria for blood donation (e.g., not pregnant within the last six weeks, never been diagnosed or have risk factors for transfusion transmitted infections such as HIV, hepatitis B virus or hepatitis C virus). Basic demographic information (age, sex, hospitalization with COVID-19) was obtained from each donor; initial diagnosis of SARS-CoV-2 and the date of diagnosis were confirmed by medical chart review. All donors provided informed consent and ∼25 mL of blood was collected in acid citrate dextrose (ACD) tubes. The samples were separated into plasma and peripheral blood mononuclear cells within 12 hours of collection. The plasma samples were immediately frozen at -80°C.

To test samples prior to the COVID-19 pandemic, stored serum specimens from an identity-unlinked HIV serosurvey conducted in 2016 among adult patients attending the Johns Hopkins Hospital Emergency Department were included (n=87). Pre-pandemic specimens were excess (i.e. discarded) samples from patients who had blood drawn for clinical purposes. (24)

Both parent studies were cross-sectional and no individual contributed multiple specimens.

### Virus and neutralization assay

The SARS–CoV-2/USA-WA1/2020 virus was obtained from BEI Resources. The virus was grown and infectious virus titrated on VeroE6TMPRSS2 cells at 33°C in infection media which is identical to the media used to grow the cells except the FBS was reduced to 2.5% as described previously for SARS-CoV-2. (25)

Plasma neutralization assays were performed essentially as described. (6, 26) SARS-CoV-2 was added to serial plasma dilutions, and the plasma-virus mixture was incubated with VeroE6TMPRSS2 cells. (27) Cytopathic effect was scored following 4% formaldehyde fixation and staining with Napthol Blue-Black, and an AUC, representing plasma neutralizing activity, was determined.

### Phagocytosis assay

ADCP was assessed as described. (11) Briefly, uptake of fluorescent RBD-conjugated microspheres by monocytic human THP-1 cells in the presence of convalescent plasma was defined by flow cytometry.

### CD16 reporter assay

FcgRIIIa ligation activity was defined as previously reported (11) as a surrogate of ADCC activity. The ability of convalescent plasma to induce luciferase expression in a Jurkat reporter cell line via ligation of FcgRIIIa was defined by culturing on RBD-coated high binding microtiter plates in the presence of convalescent plasma.

### Complement deposition assay

ADCD of heat inactivated plasma was quantified as previously (11) by incubation of RBD-conjugated multiplex assay microspheres in the presence of human complement serum and subsequent quantification of complement cascade product C3b deposition by flow cytometry.

### Luminex assessment of whole S IgG

Prefusion-stabilized, trimer-forming spike protomers (S-2P) of SARS-CoV-2 were expressed in Expi 293 cells, purified via affinity chromatography, and covalently coupled to Luminex Magplex magnetic microspheres as described. (11) Antigen specific antibodies were detected with secondary anti-IgG-PE and median flourescent intensity was measured on a FlexMap 3D array reader.

### Programmable phage display immunoprecipitation and sequencing

The design and cloning of the 56 amino acid coronavirus libraries were previously described. (10) Phage immunoprecipitation and sequencing was performed according to a previously published protocol. (28) Briefly, 0.2 μl of each plasma was individually mixed with the coronavirus phage library and immunoprecipitated using protein A and protein G coated magnetic beads. A set of 6-8 mock immunoprecipitations (no plasma input) were run on each 96 well plate. Bead washing was implemented on a Bravo liquid handling robot. Magnetic beads were resuspended in PCR master mix and subjected to thermocycling. A second PCR reaction was employed for sample barcoding. Amplicons were pooled and sequenced on an Illumina NextSeq 500 instrument.

### Statistics

Sequencing reads were mapped to library peptides using exact matching. A Bayesian hierarchical model was used to analyze the read count data and infer peptide enrichment. The read counts for each peptide were modeled as Binomial(n,p) distributions based on the total sample read counts (depth) n, and probabilities p drawn from Beta distributions. Posterior distributions for enrichment were based on 10,000 iterations derived from JAGS with slice sampling as implemented in R (using a binary yes/no indicator for enrichment in the chain), comparing the sample read counts to those observed in the mock IPs. Fold changes were reported conditional on enrichment status: fold changes for peptides with enrichment posterior probability less than 50% were reported as 1, and otherwise as the average fold change among the states where the above-mentioned binary indicator in the chain indicated enrichment (by definition, more than 5,000 iterations). Each peptide in the coronavirus VirScan library is represented by duplicate peptides, distinguishable by unique codon usage. The greater of the two fold changes of the technical replicate peptide pairs was used for downstream analyses.

Differences among virus-specific scores between sample groups were assessed via two-sided Wilcoxon rank sum tests. Wilcoxon rank sum tests were additionally used to evaluate differences in functionality of plasma with, versus without, particular peptide enrichments. Correlations between number of reactivities versus NT AUC, as well as between CoV2 and HKU1 peptides, were determined via Pearson correlation. Differences in enrichment rate between sample groups was calculated with Fisher’s exact test. When multiple tests were used, significance was determined via the Benjamini-Hochberg procedure. All analyses and visualizations were performed in R, (29) supported by R package pheatmap. (30)

### Deconvolution of VirScan data

The deconvolution algorithm to infer on-target reactivity versus off-target cross-reactivity was written in R. It was designed to determine the true targets of antibodies detected with VirScan under the assumption that reactivity (fold-change of peptide enrichment) between an antibody and its true target will exceed that of off-target cross-reactivity. First, all of the 56 amino acid peptides from SARS-CoV-2 or the endemic CoVs in the VirScan library were aligned with blastp. Peptides from the same protein of origin that had alignments with Evalue less than 100 were considered potentially cross-reactive. Fig S6 shows the peptide-peptide sequence homology map from blastp alignment of all CoV2 S, M and N peptides against all HCoV S, M and N peptides. Peptides were visualized as nodes and were colored according to the virus from which they were derived; peptides were linked if they shared alignments with an E-value less than 100. For a peptide of interest (peptide A), the reactivity was compared to the reactivities of peptides from different CoVs that aligned to peptide A (comparison peptides). Binding “preference” required the fold change of peptide A be greater than the fold changes of all comparison peptides plus a factor accounting for technical dispersion unique to each peptide pair. In order to establish the dispersion factor, we took advantage of the fact that each peptide in the library is represented in duplicate. The standard deviation of the difference between technical replicate peptides was used as a measure of expected variance in peptide reactivity, and the dispersion factor was set at 2 times this standard deviation for each peptide pair. Fig S5 contains a flow chart describing the deconvolution algorithm.

### Neutralizing titer and whole S antibody titer discordance analysis

If NT AUC predicted by a simple linear regression was greater than 40 plus 2 times the measured NT AUC, the sample was said to have discordantly High S. Similarly, if a sample’s measured NT AUC was greater than 40 plus 2 times predicted NT AUC, it was said to have discordantly Low S.

The HKU1 S CS+/CoV2 S CS-transformation was designed to normalize the error in predicted NT AUC. The log2 of the median measured NT AUC to predicted NT AUC ratio for HKU1 S CS+/CoV2 S CS-samples was -0.72, while the log2 of the median measured NT AUC to predicted NT AUC ratio for all other samples was -1.67. We normalized this ratio for the samples that were HKU1 CS positive and Cov2 CS negative to the ratio for all other samples to account for this difference.

### Study approval

Written consent was received from all study participants. The Johns Hopkins University School of Medicine Institutional Review Board reviewed and approved the sample collection and this study.

## Data Availability

Data is available upon reasonable request from the corresponding author.

## Author Contributions

W.M. designed and performed data analysis and wrote manuscript. S.H. designed and conducted experiments and edited manuscript. D.M. designed and performed data analysis and edited manuscript. A.C. processed raw data and edited manuscript. E.M.B. collected and managed samples and edited the manuscript. E.F. designed and provided reagents. I.R. designed analyses and edited the manuscript. K.L., A.R.C., H.N., S.E.B., and A.P designed and performed experiments and edited manuscript. J.A.W. managed data and samples and edited manuscript. M.Z.L performed experiments and provided reagents. T.S.B., S.E.B., A.B., S.S., D.S., and O.L. procured samples and edited the manuscript. T.C.Q. procured samples and edited the manuscript. S.E. and A.C. advised research and edited the manuscript. A.D.R. conceived project and edited manuscript. M.E.A. supervised research and edited manuscript. S.J.E. oversaw reagent development, provided reagents, and edited the manuscript. M.R. supervised the research and wrote manuscript. A.A.R.T. conceived project and edited manuscript. H.B.L. conceived project, supervised research, and wrote manuscript.

## Acknowledgements

This study was made possible by grants from: the Johns Hopkins COVID-19 Research Response Program, National Institute of General Medical Sciences (NIGMS) grant R01GM136724 and a National Institute of Allergy and Infectious Diseases (NIAID) grant U24AI118633 (H.B.L.); NIAID R01AI120938, R01AI120938S1 and R01AI128779 (A.A.R.T); NIH grants AI052733, AI15207 and HL059842 (A.C.); in part by the Division of Intramural Research, NIAID, NIH (O.L., A.R., T.Q.); National Heart Lung and Blood Institute 1K23HL151826-01 (E.M.B); Bloomberg Philanthropies (A.C.); Department of Defense W911QY2090012 (D.S.); NIH Center of Excellence for Influenza Research and Surveillance grant HHSN272201400007C (A.P.); the MassCPR and VOVRN (S.J.E); and HIV Prevention Trials Network grant UM1AI068613 (S.E.) We are grateful to the National Institute of Infectious Diseases, Japan, for providing VeroE6TMPRSS2 cells and acknowledge the Centers for Disease Control and Prevention, BEI Resources, NIAID, NIH for providing SARS-related coronavirus 2, isolate USA-WA1/2020, NR-5228.

---

**Figure S1.**
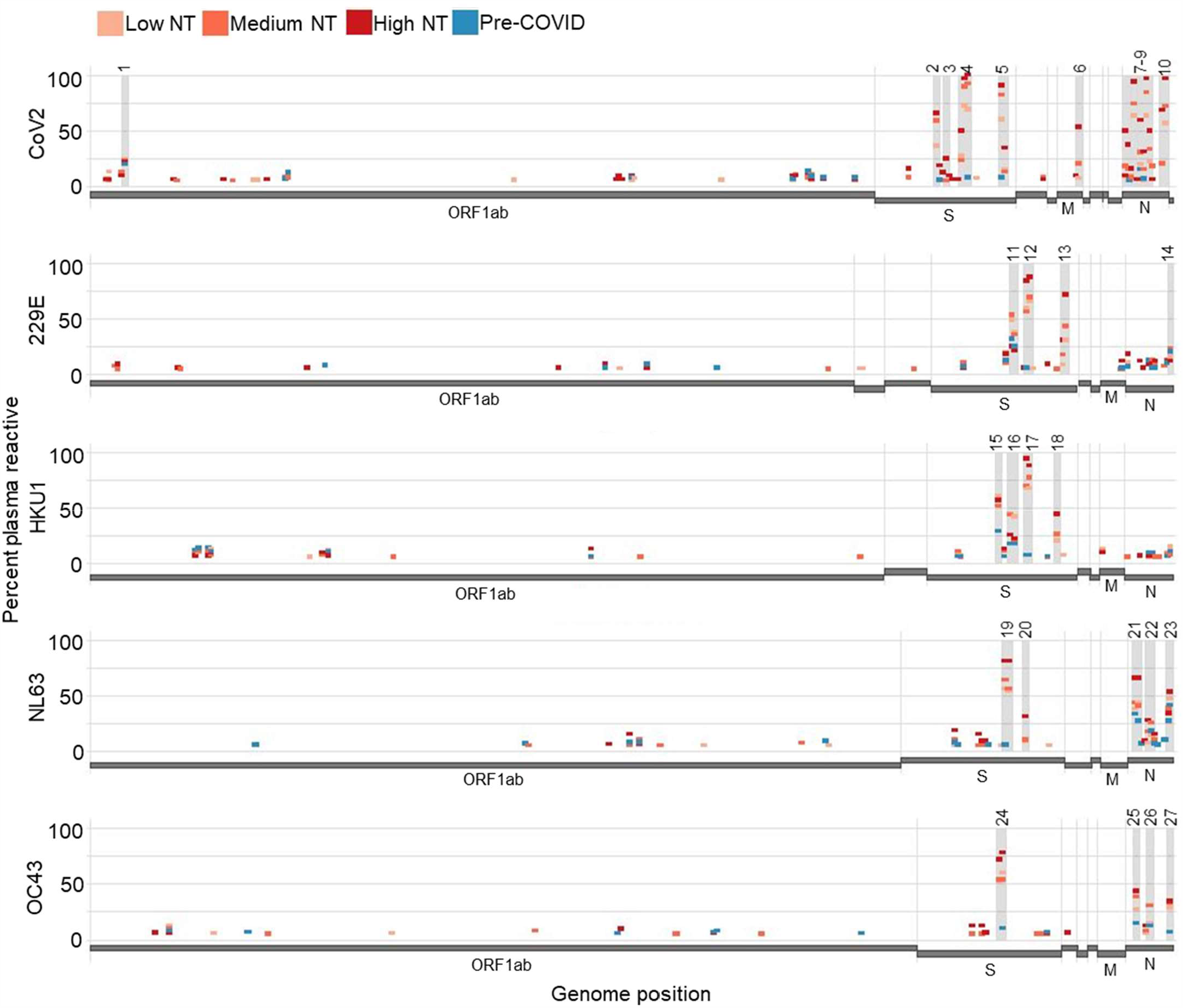
Whole genome reactivity plots. Antibody reactivity plots analogous to those in **Fig 1** were created that include the poorly reactive ORF1.

**Figure S2.**
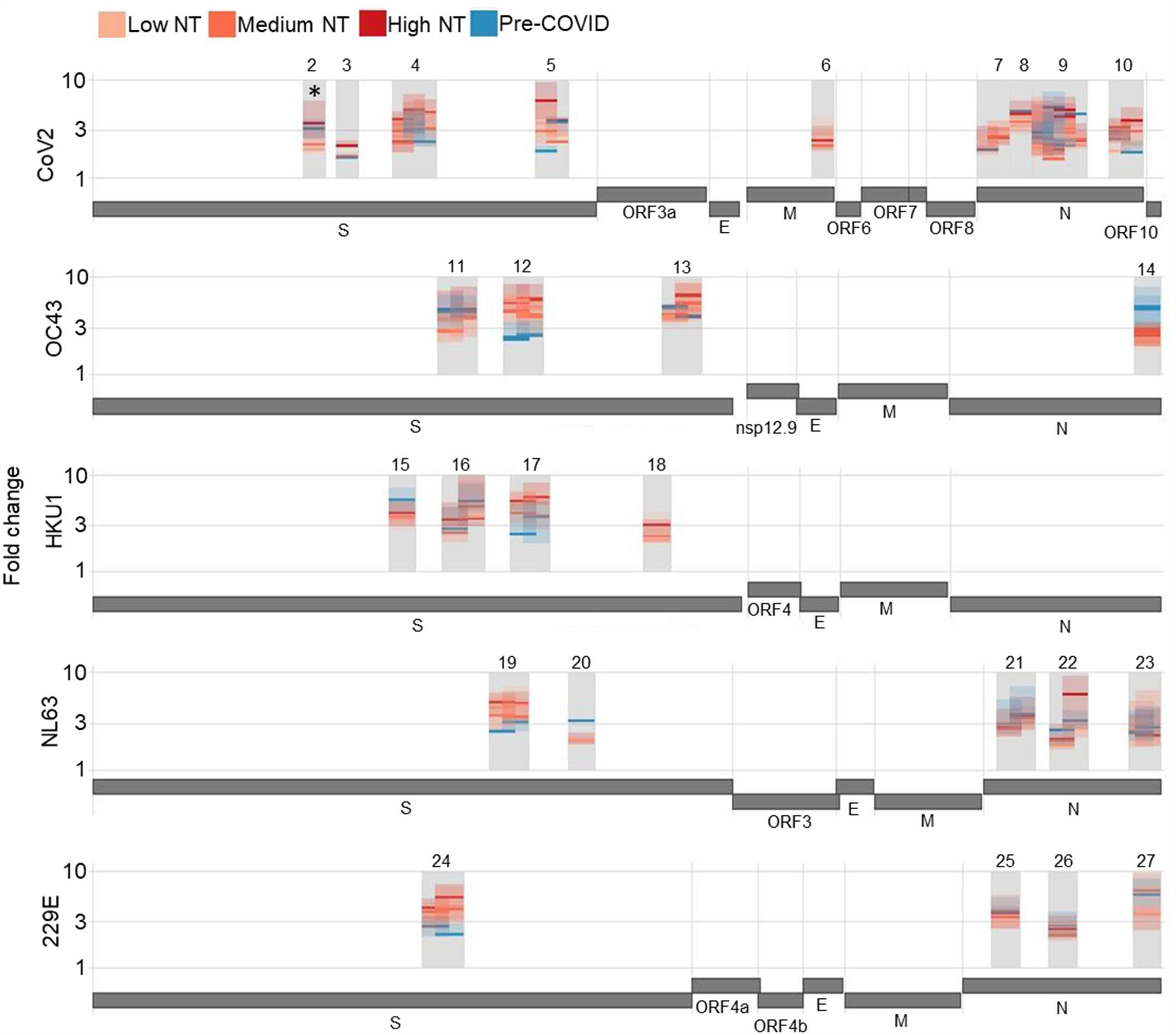
Magnitude of peptide reactivities does not distinguish plasma functionality. The median and interquartile range of antibody reactivity for each sample group is plotted for each immunodominant peptide. One CoV2 S immunodominant peptide (residues 533-588) indicated by asterisk show greater magnitude reactivities in High NT plasma compared to Medium NT or Low NT plasma (Wilcox test vs Medium NT p=0.014, vs Low NT p<0.001).

**Figure S3.**
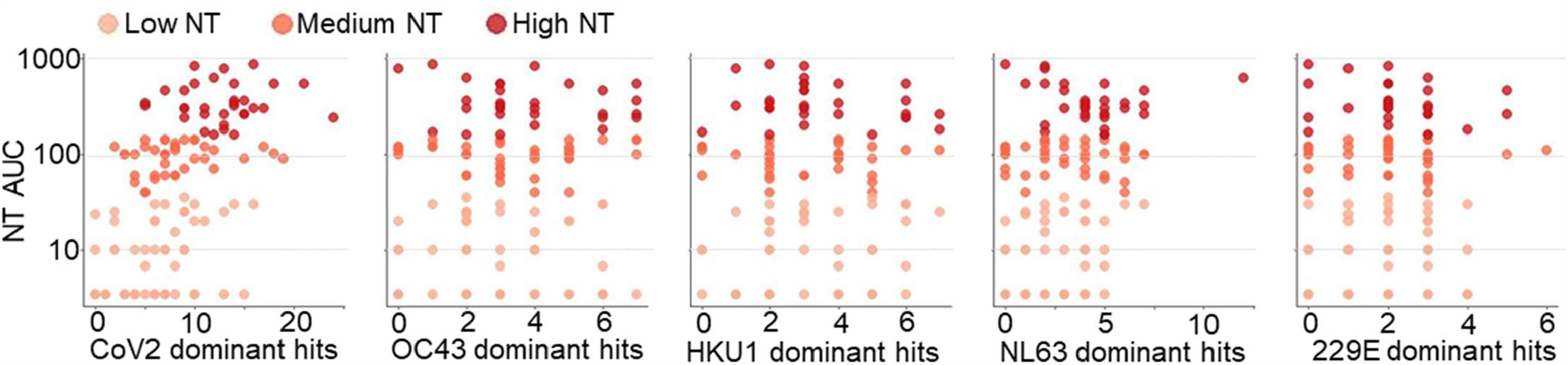
Polyclonality of antibody responses to CoV2 and NL63 immunodominant regions is associated with increased NT AUC. Number of reactive peptides from immunodominant regions of each coronavirus was compared to NT AUC. Polyclonal responses to CoV2 and NL63 correlate with increase NT AUC (Pearson’s correlation, CoV2 p< 10^−8^, R=.49; NL63 p=0.02, R=.21).

**Table S1.**
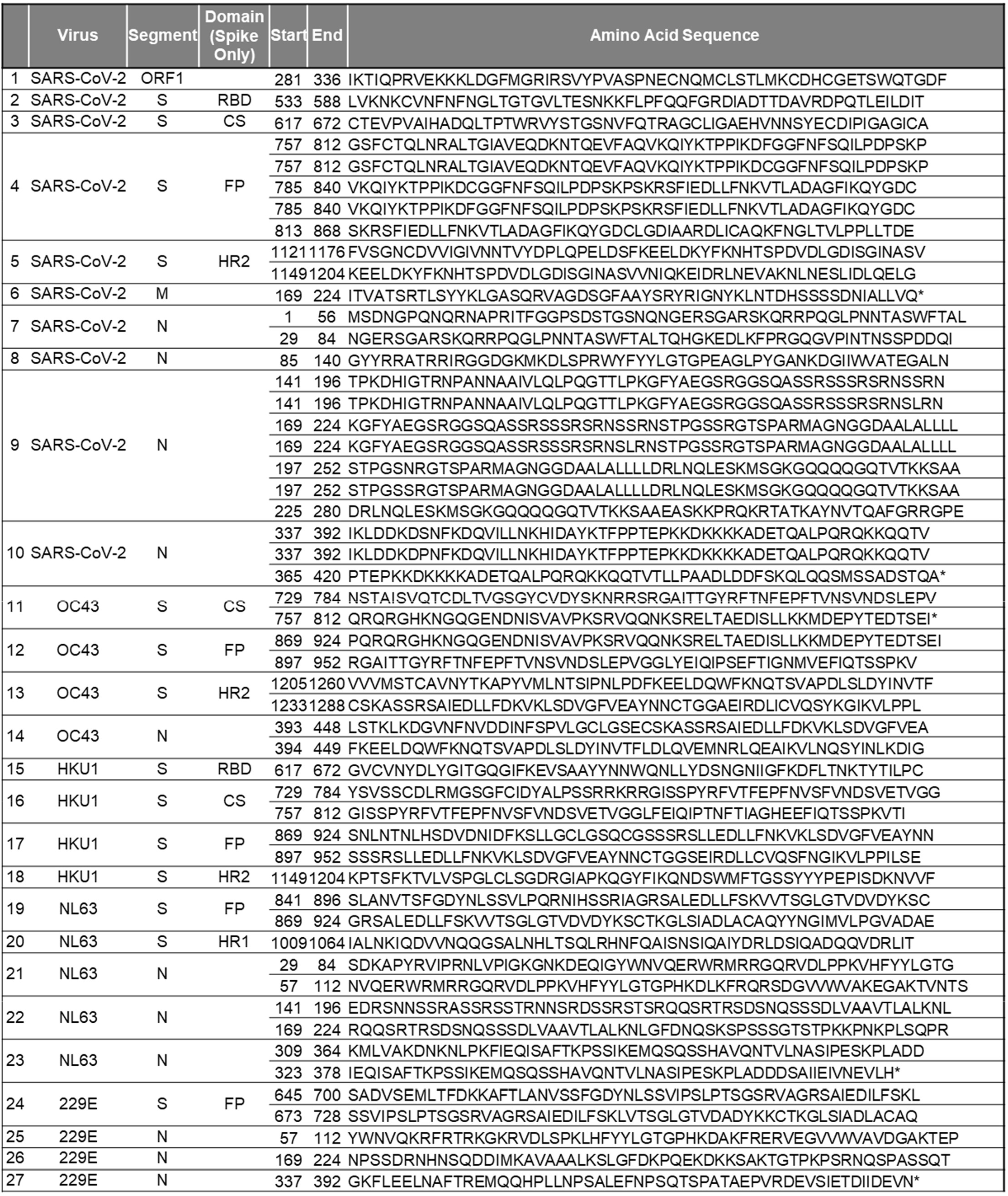
Amino acid sequences of immunodominant CoV2 and HCoV peptides.

**Figure S4.**
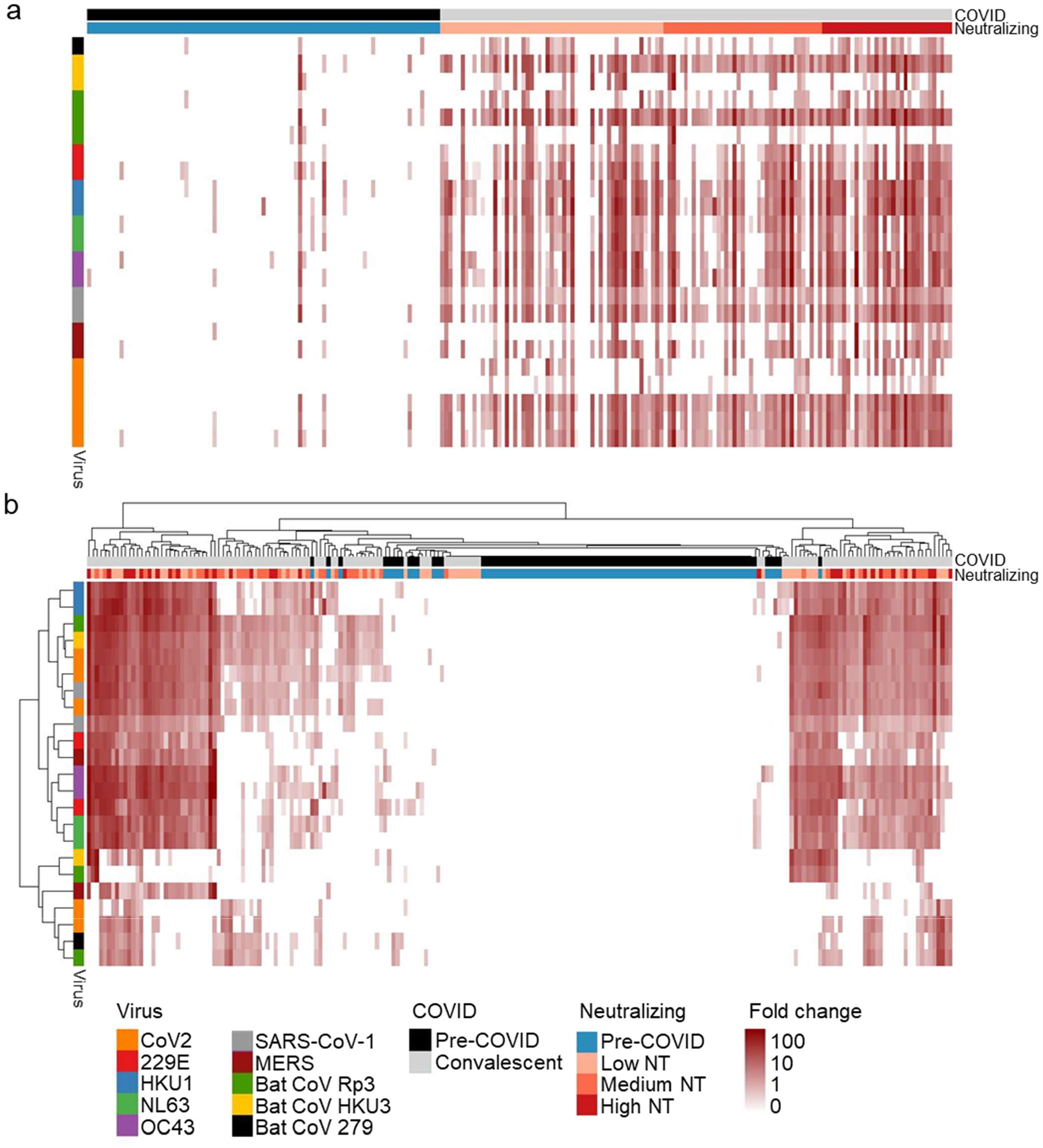
Pan-coronavirus fusion peptide antibody reactivity. Antibody binding of all study samples to all CoV FP peptides are shown in the form of a heatmap ordered by virus and sample group (**a**) and as a clustered heatmap (**b**). The FP of all CoVs represented in the VirScan library showed sequence homology to the dominant CoV2 FP peptides; reactivity was detected against every CoV FP.

**Figure S5.**
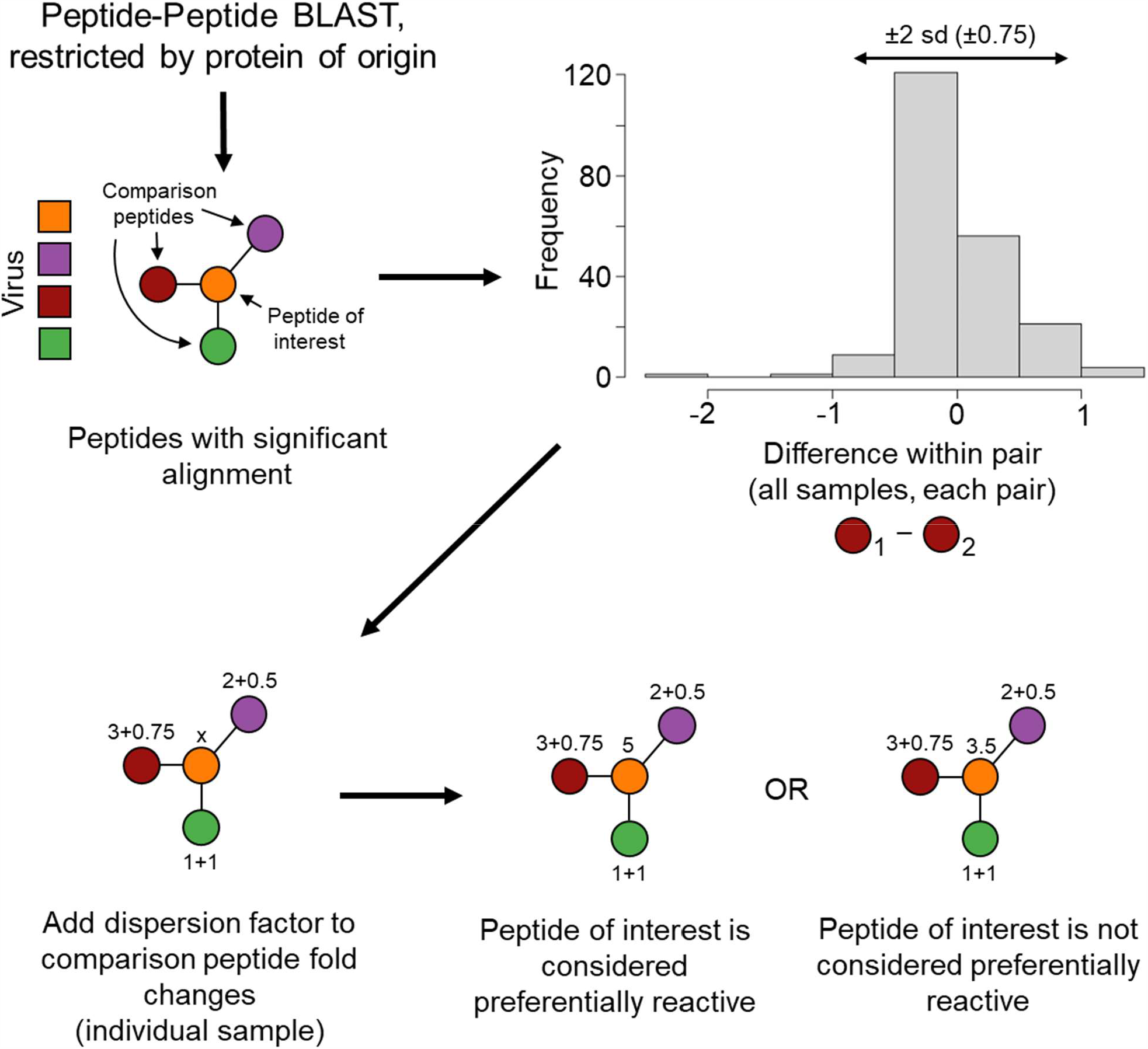
Deconvolution algorithm schematic. Peptides from different viruses of origin and the same protein of origin underwent peptide-peptide blastp. If peptides showed significant alignment (evalue<100), they were considered potentially cross-reactive. By taking advantage of the duplicate representation of each peptide in the library, a measure of expected technical dispersion was calculated. If a target peptide displayed enrichment greater than any comparison peptide plus the factor for technical dispersion, it was considered an on-target reactivity.

**Table S2.** Deconvoluted immunodominant coronavirus peptides and their functional correlates. The frequency of enrichment of 52 immunodominant peptides among each sample group is shown post deconvolution. The percentage of samples with a specific reactivity is shown (red shading). Associations with plasma functionality were defined by dichotomizing all plasma by presence or absence of each particular reactivity followed by a two sided Wilcox test. The negative log transformed p values are shown (green shading).

|  | Virus | Segment | Domain<br>(Spike Only) | Start | End | Percent Samples Enriched |  |  |  | Association with Phenotype (-log(p)) |  |  |  |
| --- | --- | --- | --- | --- | --- | --- | --- | --- | --- | --- | --- | --- | --- |
|  |  |  |  |  |  | Pre-COVID<br>n=87 | Low NT<br>n=55 | Medium<br>NT n=39 | High NT<br>n=32 | NT AUC | ADCP | ADCC | ADCD |
| 1 | SARS-CoV-2 | ORF1 |  | 281 | 336 | 8 | 7.3 | 10.3 | 9.4 | 0.3 | 0.2 | 0.3 | 0.2 |
| 2 | SARS-CoV-2 | S | RBD | 533 | 588 | 1.1 | 1.8 | 10.3 | 18.8 | 2.6 | 4.4 | 4.7 | 3.7 |
| 3 | SARS-CoV-2 | S | CS | 617 | 672 | 1.1 | 0 | 5.1 | 25 | 3.5 | 4.5 | 3.6 | 1 |
| 4 | SARS-CoV-2 | S | FP | 757 | 812 | 0 | 9.1 | 7.7 | 9.4 | 0 | 0.3 | 0.4 | 0.2 |
|  |  |  |  | 757 | 812 | 0 | 20 | 17.9 | 34.4 | 1 | 2.4 | 2.3 | 3.1 |
|  |  |  |  | 785 | 840 | 1.1 | 0 | 2.6 | 6.3 | 1 | 0.7 | 1 | 1.2 |
|  |  |  |  | 785 | 840 | 0 | 1.8 | 10.3 | 9.4 | 1.4 | 1.4 | 0.7 | 1.6 |
|  |  |  |  | 813 | 868 | 0 | 0 | 5.1 | 3.1 | 0.6 | 1.1 | 0.5 | 0.1 |
| 5 | SARS-CoV-2 | S | HR2 | 1121 | 1176 | 0 | 20 | 23.1 | 40.6 | 1.4 | 2.7 | 1 | 1.8 |
|  |  |  |  | 1149 | 1204 | 1.1 | 5.5 | 0 | 0 | 1.1 | 0.5 | 0.2 | 0.2 |
| 6 | SARS-CoV-2 | M |  | 169 | 224 | 0 | 7.3 | 20.5 | 50 | 4.7 | 5.8 | 6.7 | 1.5 |
| 7 | SARS-CoV-2 | N |  | 1 | 56 | 1.1 | 9.1 | 17.9 | 50 | 4.8 | 4.7 | 6.4 | 3.5 |
|  |  |  |  | 29 | 84 | 0 | 5.5 | 15.4 | 37.5 | 4.4 | 3.5 | 4.4 | 2.1 |
| 8 | SARS-CoV-2 | N |  | 85 | 140 | 0 | 34.5 | 48.7 | 46.9 | 1.5 | 4.7 | 1.6 | 0.6 |
| 9 | SARS-CoV-2 | N |  | 141 | 196 | 11.5 | 18.2 | 25.6 | 43.8 | 1.5 | 1 | 3.2 | 3.4 |
|  |  |  |  | 141 | 196 | 14.9 | 20 | 30.8 | 59.4 | 2.5 | 1.9 | 3.2 | 3.3 |
|  |  |  |  | 169 | 224 | 2.3 | 5.5 | 2.6 | 12.5 | 0.7 | 0.1 | 1.4 | 1 |
|  |  |  |  | 169 | 224 | 5.7 | 3.6 | 5.1 | 18.8 | 1.5 | 1 | 1.3 | 2 |
|  |  |  |  | 197 | 252 | 2.3 | 49.1 | 76.9 | 96.9 | 4.9 | 3.9 | 3.4 | 2.7 |
|  |  |  |  | 197 | 252 | 3.4 | 47.3 | 64.1 | 93.8 | 7.1 | 6.8 | 3.7 | 2.4 |
|  |  |  |  | 225 | 280 | 1.1 | 20 | 33.3 | 40.6 | 1.7 | 3 | 4.2 | 1.8 |
| 10 | SARS-CoV-2 | N |  | 337 | 392 | 2.3 | 20 | 17.9 | 65.6 | 4.3 | 4.1 | 5.4 | 3.2 |
|  |  |  |  | 337 | 392 | 2.3 | 16.4 | 17.9 | 56.3 | 4.6 | 5 | 5.9 | 3.1 |
|  |  |  |  | 365 | 420 | 3.4 | 56.4 | 71.8 | 96.9 | 4.3 | 4.3 | 4.1 | 0.7 |
| 11 | OC43 | S | CS | 729 | 784 | 14.9 | 7.3 | 7.7 | 9.4 | 0 | 0.1 | 0.3 | 0.2 |
|  |  |  |  | 757 | 812 | 12.6 | 5.5 | 5.1 | 6.3 | 0.1 | 0.3 | 0 | 0.6 |
| 12 | OC43 | S | FP | 869 | 924 | 0 | 0 | 0 | 6.3 | 1.3 | 0.5 | 0.3 | 0.5 |
|  |  |  |  | 897 | 952 | 0 | 0 | 2.6 | 6.3 | 1.4 | 1.1 | 0.9 | 1 |
| 13 | OC43 | S | HR2 | 1205 | 1260 | 1.1 | 0 | 0 | 0 | 0 | 0 | 0 | 0 |
|  |  |  |  | 1233 | 1288 | 1.1 | 9.1 | 10.3 | 6.3 | 0 | 0 | 0.3 | 0.5 |
| 14 | OC43 | N |  | 393 | 448 | 20.7 | 9.1 | 15.4 | 9.4 | 0.2 | 0 | 0 | 0.4 |
|  |  |  |  | 394 | 449 | 19.5 | 10.9 | 17.9 | 9.4 | 0.2 | 0 | 0.2 | 0.1 |
| 15 | HKU1 | S | RBD | 617 | 672 | 28.7 | 50.9 | 35.9 | 40.6 | 1 | 1 | 0.6 | 0.1 |
| 16 | HKU1 | S | CS | 729 | 784 | 1.1 | 7.3 | 2.6 | 6.3 | 0.1 | 0 | 0 | 0 |
|  |  |  |  | 757 | 812 | 8 | 10.9 | 2.6 | 6.3 | 0.7 | 0.2 | 0.3 | 0.1 |
| 17 | HKU1 | S | FP | 869 | 924 | 1.1 | 3.6 | 0 | 6.3 | 0.2 | 0.3 | 0.3 | 0.2 |
|  |  |  |  | 897 | 952 | 0 | 12.7 | 2.6 | 12.5 | 0 | 0.5 | 1.3 | 0.6 |
| 18 | HKU1 | S | HR2 | 1149 | 1204 | 0 | 20 | 23.1 | 43.8 | 1.6 | 2.4 | 3.1 | 4.5 |
| 19 | NL63 | S | FP | 841 | 896 | 0 | 0 | 0 | 3.1 | 0.7 | 0.9 | 0.8 | 0.6 |
|  |  |  |  | 869 | 924 | 0 | 0 | 0 | 0 | 0 | 0 | 0 | 0 |
| 20 | NL63 | S | HR1 | 1009 | 1064 | 0 | 5.5 | 5.1 | 18.8 | 1.3 | 0.3 | 0.3 | 0.3 |
| 21 | NL63 | N |  | 29 | 84 | 13.8 | 5.5 | 2.6 | 0 | 1.3 | 1.2 | 0 | 0.6 |
|  |  |  |  | 57 | 112 | 16.1 | 7.3 | 12.8 | 9.4 | 0.1 | 0.6 | 0.3 | 0.8 |
| 22 | NL63 | N |  | 141 | 196 | 1.1 | 3.6 | 2.6 | 12.5 | 0.8 | 0.3 | 1.1 | 0.8 |
|  |  |  |  | 169 | 224 | 17.2 | 9.1 | 15.4 | 15.6 | 0.7 | 0.1 | 0.1 | 0.2 |
| 23 | NL63 | N |  | 309 | 364 | 8 | 3.6 | 7.7 | 3.1 | 0.2 | 0.2 | 0.1 | 0 |
|  |  |  |  | 323 | 378 | 14.9 | 9.1 | 15.4 | 12.5 | 0 | 0.2 | 0.1 | 0.4 |
| 24 | 229E | S | FP | 645 | 700 | 0 | 0 | 0 | 0 | 0 | 0 | 0 | 0 |
|  |  |  |  | 673 | 728 | 0 | 1.8 | 0 | 0 | 0.2 | 0.2 | 1 | 0.5 |
| 25 | 229E | N |  | 57 | 112 | 5.7 | 10.9 | 5.1 | 0 | 1.8 | 1.1 | 1.2 | 0.5 |
| 26 | 229E | N |  | 169 | 224 | 5.7 | 5.5 | 5.1 | 0 | 0.5 | 0.3 | 0.5 | 0.3 |
| 27 | 229E | N |  | 337 | 392 | 4.6 | 10.9 | 15.4 | 9.4 | 0.4 | 0.1 | 0.2 | 0.4 |

**Figure S6.**
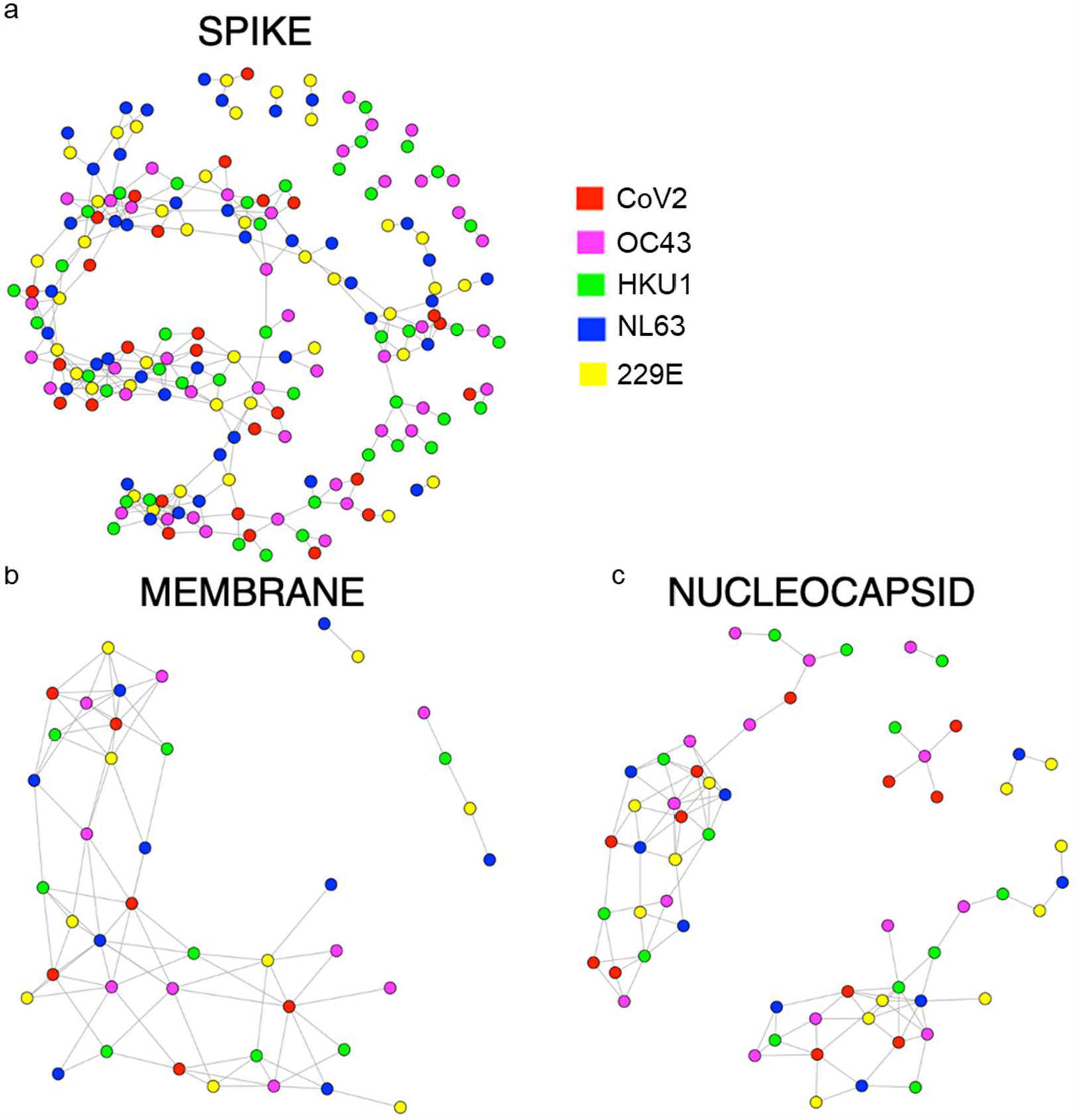
Visualization of CoV2 and HCoV peptide sequence homologies. Network graphs show sequence homologies among the S (**a**), M (**b**) and N (**c**) peptides between CoV2 and each HCoV. Nodes (peptides) are colored by their corresponding virus. Peptides are linked by an edge if they share blastp sequence similarity. Only homologies among peptides from different viruses are shown for simplicity.

